# MODELING COVID19 IN INDIA (Mar 3-May 7, 2020): HOW FLAT IS FLAT, AND OTHER HARD FACTS

**DOI:** 10.1101/2020.05.11.20097865

**Authors:** Smarajit Dey

## Abstract

A time-series model was developed for *Number of Total Infected Cases in India*, using data from *Mar 3 to May 7, 2020*. Two models developed in the early phases were discarded when they lost statistical validity, The third, current, model is a 3^rd^-degree polynomial that has remained *stable over the last 30 days (since Apr 8), with R^2^ > 0.998 consistently*. This model is used to forecast Total Covid cases, after cautionary discussion of triggers that would invalidate the model. The purpose of all forecasts in the study is to provide *a comparator* to evaluate policy initiatives to control the pandemic - the forecasts are not objectives by themselves. Actual observations less than forecasts mean successful policy interventions. Figures of *Doubling Time*, Fatality Rate and Recovery Rate used by authorities are questioned. Elongation of doubling rates is inherent in the model, and worthy of mention only when the time actually exceeds what the model predicts. The popular *Fatality Rate* and *Recovery Rate* metrics are shown to be illogical. The study defines two terms *Ongoing Fatality Rate (OFR)*, and *Ongoing Recovery Rate (ORR)* and determines these currently to be *~9% and ~76%* respectively in India. Over time, OFR will decline to the eventual Case Fatality Rate (CFR), while ORR will eventually climb to (1-CFR). There is no statistical basis to assume eventual Indian CFR, and China’s 5.5% CFR is used as a proxy. *Using these metrics, the current model forecasts by May-end, >150000 Total Infected, ~5000 Deaths and >85000 Active Cases*. There is no *pull-back* evident in the current model in the foreseeable future, and cases continue to rise at progressively slower rates. Subject to usual caveats, the model is used to forecast till Sep 15. *The study argues that Indian hospital infrastructure is reasonably ready to handle Active Cases as predicted for Sept 15* – in that sense, the curve is “flat enough”. *However, the curve is NOT flat enough with respect to fatalities - nearly 100000 by Sept 15*. Setting an arbitrary limit that Total Deaths must be within 50,000 by Sept 15, the study retrofits a model that shows what the desired growth of Covid19 cases should be. *It is seen that overall doubling time of 38 days is required in period June 1 to Sep 15, if deaths are to remain below 50000*.

---

The phenomenon under observation is the Total number of Covid19 Cases (total infected) in India, day-wise from March 3^rd^ 2020 till May 7th. Visualise a graph depicting this number over time. *Mathematical modelling* attempts to discover underlying patterns in this graph. *Mathematical models* are equation(s) that express the underlying patterns. In this case, our model provides an estimate of Total Covid Cases on a given date. For dates past, the estimate can be compared against the actual number observed. Intuitively, if the deviations between estimates (by model) and actuals (observed) are small, then it is a model with robust predictability, which can be used to forecast numbers for the future. The model’s robustness of predictability, usually denoted by **R^2^**, is a figure between 0 and 1 (1 denoting best fit between model and observed data). All our models have had exceptionally high R^2^.

A model does not *determine* or *influence* the real-life progression of the observed data – it ***reflects*** it. *Real-life events* influence the progression. In the case of Total Covid Cases, disruptive events can be planned initiatives (lockdown, containment, mass movements), or unplanned reckless social behaviour. It is known that the effects of such events show up in observed Total Cases with a lag of 1 to 2 weeks. The change in the nature of progression *should show up* in the output of the mathematical model as well. This does not happen automatically – what happens is that estimates thrown-up by the model begin deviating significantly from observed data. This is the signal that the underlying pattern has changed, and so the model must be revised to reflect the new underlying pattern. Thus, over the life of this pandemic, we expect to see different models in different phases, as the real-life nature of progression changes.

**Conversely, when a model continues to present a close fit between model-output and actuals (high R^2^) for a long time, then it means that the model is well-constructed and robust enough** to absorb small deviations in observed data. *Such a model, stable over time, gives confidence when used to make forecasts*. **The current model in our study has remained unchanged for the last 30 days with R^2^ >=0.998 throughout** (reporting date 8^th^ May, with data upto 7^th^ May).

*Assuming that the model continues unchanged*, we forecast numbers of Covid19 cases in India. Separately, we also examine Fatalities, Recoveries and Active Cases, and derive conclusions. The study is not important for the forecasts it makes. The study is directionally important to identify trends and trajectories. **But most of all the study is a comparator to measure the consequences of policy action. If actual cases are lower than forecasted numbers, then we are doing well**.

## A) METHODOLOGY OF MODELING

Data on Daily Infected Cases (Fresh Cases), Daily Deaths, Daily Recoveries and Daily Active Cases have been consistently sourced from the daily 8am update of the MoHFM**^(1)^** from **3^rd^ March 2020 (Day 1)** – data reported at 8am 4^th^ Mar – from the time the 4^th^ case of Covid19 was recorded. [Initial 3 diagnosed/recovered in Jan/Feb.] A 2-day Moving Average (2dMA) of Fresh Cases was done to smoothen random delays in Test sample processing, Results Announcement, etc. The daily 2dMAs were cumulated to give the *daily Total Cases*. Modelling commenced from Day 13 (15^th^ March), when the total cumulated 2dMAs (Total Infected Cases) reached 100.

Three models have been successively developed so far to depict the changing pattern of Total Cases progression. Table 1 below provides details of the three models.

**Table 1:** Details of 3 Models Used to Depict Total Cases, with Phase 3 remaining current.

| Phase | Valid over the time period... |  |  | Created On | Nature of Model | R-Squared |  |  |  |
| --- | --- | --- | --- | --- | --- | --- | --- | --- | --- |
|  | Days | Dates | Duration |  |  | Observed | Mean | Max | Min |
| One | D1-D22 | Mar 3 - 24 | 22 days | Mar 15 (D13) | 3<sup>rd</sup> degree Polynomial | D13-D22 | 0.9879 | 0.995 | 0.977 |
| Two | D23-D36 | Mar 25 - Apr 7 | 14 days | Apr 4 (D33) | Exponential | D23-D36 | 0.9962 | 0.999 | 0.990 |
| Three | D37-D66+ | Apr 8 - May 7++ | 30+ days | Apr 12 (D41) | 3<sup>rd</sup> degree Polynomial | D37-D66+ | 0.9983 | 0.999 | 0.994 |

Models of Phase 1 and Phase 2 are shown in Figs 1 and 2, where the blue lines are observed cases and the red lines the values predicted by the model. Contrary to intuitive understanding of the nature of a pandemic, the initial model in Fig 1 was not an exponential curve (R^2^ only 0.85), but a 3^rd^ degree polynomial. Fig 1 also shows that the initial trajectory changed significantly from Day 22 (Mar 24), when the pandemic took hold, and this trend accelerated from Day 31 (Apr 2–sudden spike of many cases), confirming that this model was no longer usable. The model that matched observed data from Day 23 (Mar 25) to Day 36 (Apr 7) was an exponential model. It can be seen from Fig 2 that the actual observed cases were less than predicted ones from Day 37 (Apr 8) onwards. One can speculate that the effects of the lockdown were showing up with the expected lag 7-14 days. It became necessary to use a fresh model to depict the situation from Day 37 onwards, and this model less-steep than the previous one.

**Fig 1:**
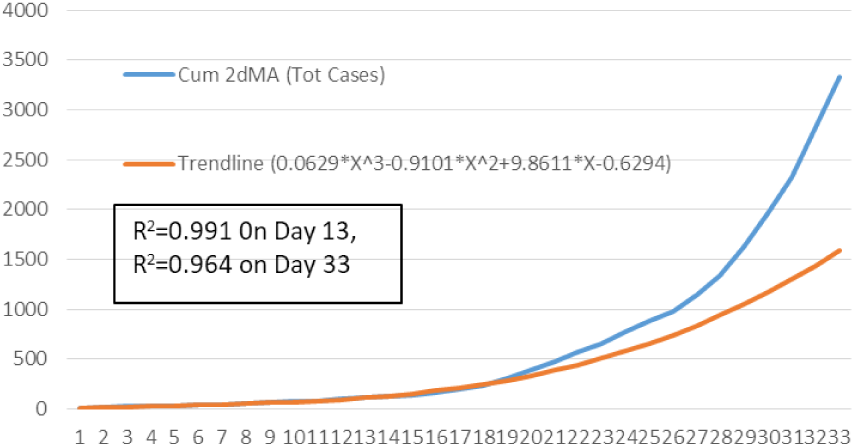
Polynomial Fit for Day 1 – Day 22 (Mar 3 - 24)

**Fig 2:**
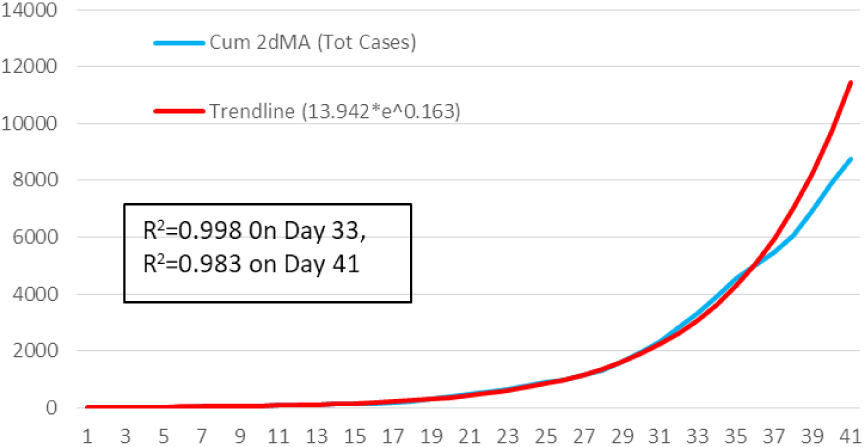
Exponential Fit for D23-D36 (Mar 25 – Apr 7)

The 3^rd^ degree polynomial that modelled Total Cases from Day 37 (Apr 8) onwards, shown in Fig 3 has remained remarkably resilient and robust throughout the last 30 days (May 7), as can be seen from Table 1 above, showing mean R^2^ over 30 days to be 0.9983. It continues to remain stable and shows no visible deviations.

**Fig 3:**
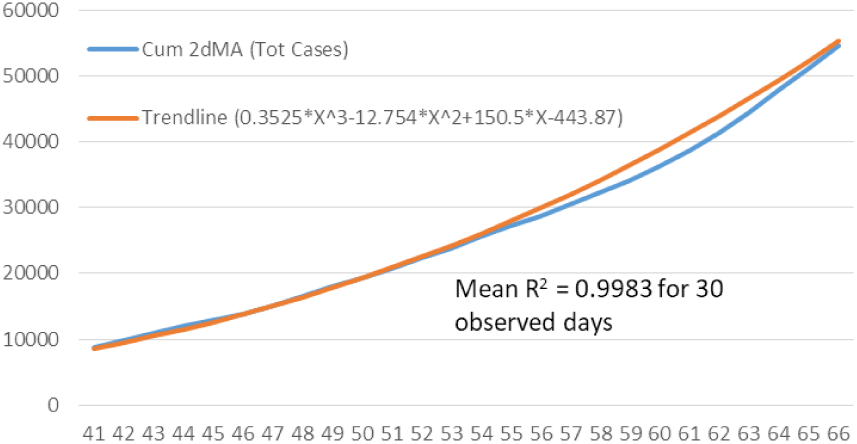
Polynomial Fit for D37 – D66+ (Apr 8 – May 7 & continuing)

Fig 4 summarises the 3 phase-wise models in one graph, red line being the observed cases.

**Fig 4:**
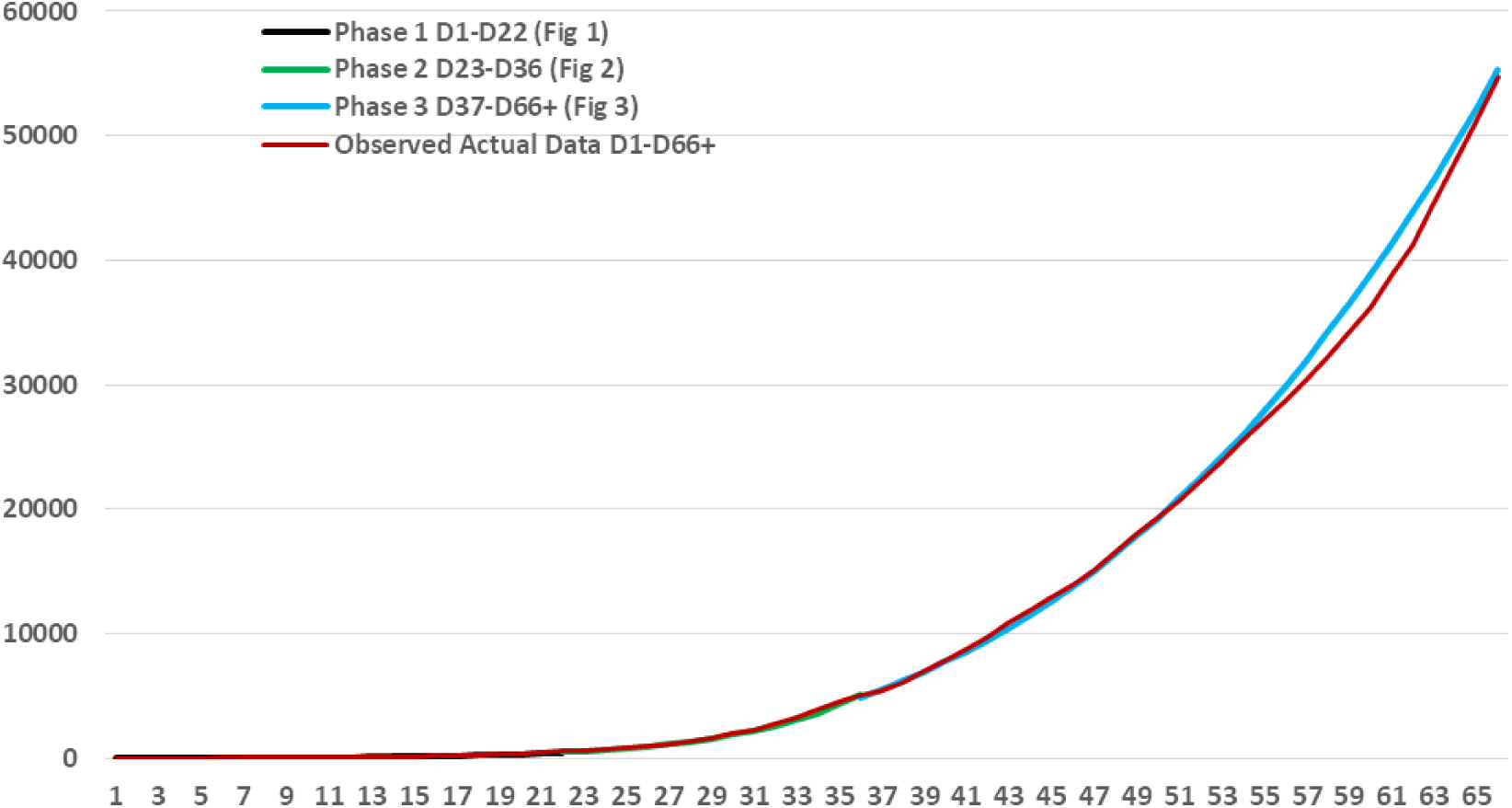
3 Phase-wise Models depict the Pandemic Mar 3 – May 7 (D1-D66)

**When and how will the model be updated?** When real-life events are significant enough to change the nature of the growth curve, this will show up as actual numbers of Covid Infections deviating from the forecasts. As in the earlier phases of the pandemic, when the deviations lead to a fall in R^2^ (robustness of predictability) – typically R^2^ <0.98 – a fresh model will be fitted spanning the period when deviant observations started. A significant event that can alter the shape of the curve can happen any time, or may have already happened and we are in the lag period. Whenever it does, the model needs to be updated. Some models may be short-lived, actuals soon beginning to deviate again. The update is then repeated afresh. When a model is stable - high R^2^ for at least 14 observations – it can be used as a basis for making further forecasts.

## B) EXPLORING TOTAL INFECTED CASES

That the current model has remained valid for 30+ days means that newsworthy events (of spikes and troughs) have not been statistically significant. In plain English, they have “cancelled out” and been “absorbed” within the predictive ability of the model. This means that without a disruption greater than in the last 29 days, this graph **would continue *on its own* pattern and trajectory – curves, lines, plateaus or whatever**. That this graph would continue on its own trajectory unless interrupted by a significant-enough intervention, ***makes it an important comparator***. For example, “highest ever cases on a day” is not a shock if the graph were forecasting a similar number. Similarly, a particular containment strategy can only be claimed to have worked if the shape of the graph changes and reaches a point lower than where *it would have naturally gone*.

Of course, this graph *will eventually* change shape, and a new model will be required for the new pattern. Significant-enough events that could make this change include medical events (vaccine, cure or acquired immunity), climatic triggers, other acts-of-God, *and of course, planned policy interventions*. For example, the strategy of actively testing large numbers, tracing and quarantining, may shortly lead to a reduction of numbers (say, if all prospects traced and isolated). The relaxations during Lockdown 3.0, planned inter-state migrations and further relaxations are other policy interventions that may change the change the shape of the graph. The last set of interventions have the potential to make the pattern steeper than at present.

### The Misrepresentation of Doubling Time

We hear regular announcements on **“doubling time”** of Total Infected Cases, and how this time has increased (to a figure of 12 days, currently). *The joy that accompanies such announcements is, unfortunately, totally misplaced*. **Increase of the doubling time is an inherent part of the pattern of the model of virus spread**. For the graph remaining unchanged, the current doubling time of 12 days, would automatically increase to 16 days by the end of May, and to 19 days by mid-June. **This reiterates one of the most important arguments of this study – that a mathematical model is *a comparator***, **and *successes occur only when the outcomes are lower than forecasts by the model***. Conclusions from this study show later that doubling time must soon rise to 38+ days for the pandemic to be under control.

### Near-Term Forecasts

A standard disclaimer that must accompany all uses of this model for forecasting is to repeat that disruptive events on-ground can/will change the model with some lag, that near-term forecasts are more plausible than longer-term ones (statistical errors of forecasting multiply for longer ones), and that even small and acceptable statistical off-targets may have huge physical implications – a 3% forecasting error, statistically acceptable, may involve 3000 human beings. Any forecast always assumes that the model remains stable.

Subject to this disclaimer, the near-term forecasts by this model are that in the weeks 4^th^-10^th^ May, 11^th^-17^th^ May, 18^th^-24^th^ May and 25^th^-31^st^ May, **the fresh cases added will exceed, respectively, 22000, 25000, 33000 and 38000. Thus, by May-end, the total number of Covid19 cases recorded would exceed 150,000**

### Long Term Issues

*The current model of Total Infected Cases has no inherent “pull-back” element which can lower its trajectory in the foreseeable future*. That is, its trajectory will keep growing at progressively slower speeds. The import of this statement is that there is no point in the foreseeable future where growth of new cases will zeroise. Strictly as per the current model, ***the country would cross a million recorded cases by early-August***.

## C) EXPLORING FATALITIES

**Current announced measures of fatality rates are flawed** because they do not reflect the disease progression. These measures divide the number of recorded deaths by the number of recorded cases to arrive the Fatality rate. This measure is incorrect.

Disease progression of Covid19 has been extensively studied. The report by Lord**^(2)^** (April 2020) concludes that the mean time from diagnosis to death for non-survivors is 18.5 days. The same report, computes mean time from diagnosis-to-cure as 17 days, and average ICU stay, for those who need it, as 15 days.

For our purpose, we will take the mean time diagnosis-to-death is 15 days. Any measure of fatality rate must naturally against the cohort who were patients at the same time. *In other words, if average time diagnosis-to-death is 15 days, the number of deaths occurring today must be compared against those who were patients 15 days ago*. We define a term **Ongoing Fatality Rate (OFR)** – *OFR on any day is the cumulative number of deaths divided by the total infected cases 15 days ago*. This is a more realistic approximation of fatality rate. To appreciate this better, visualise how the current (flawed) computation of Fatality Rate will change as Fresh Cases go down and become zero, though a few hospital patients still pass away – Fatality Rate will *increase* and that is illogical. Our definition of OFR formalises the fact that death-rate cannot be measured out of those who joined as patients within the last 15 days, because they have yet to live out their disease process.

Three further observations on OFR:

- We have defined the denominator to be Infected Cases on D-15. Since cases are growing daily, it is clear that Total Infected Cases on a later date would be higher (denominator would be larger), and consequently OFR smaller. Conversely, using Total Cases on a date earlier than 15 days would make the OFR higher. Our use of mean time diagnosis-to-death as 15 days, rather than 18.5 days reported literature, is an optimistic view of Fatality Rates.
- While OFR is our own creation, Case Fatality Rate (CFR) is a known statistic, defined as *Total* Deaths divided by *Total* Infected. Clearly, authentic CFR for an epidemic is known at the *end-point*, when fresh cases have stopped some days ago, and all active patients have recovered or expired. Since no country has reached this end-point, **all Fatality rates being reported are ongoing rates, however they may be defined**.
- Conceptually, until testing and new-case identification reaches steady-state, OFR will continue to decline as more and more cases get rapidly added. In due course, OFR *should stabilise close to the eventual CFR*.

The **Ongoing Fatality Rate (OFR) in India** has dropped sharply from ~40% in early-April to ~9% now, as testing has speeded up. It has remained steady between 8.73% and 9.71% over D57 to D65 (7^th^ May, last data date), **averaging 8.93%** over last 6 days, and declining. Since the inception of the pandemic, 8.73% has been the lowest value of OFR.

There is no way to predict where OFR in India will reach – that is, what will be the eventual Fatality Rate. Total deaths divided by total infected patients worldwide on 6^th^ May, stands at 7.02%**^(3)^**. By country, these rates vary widely, between 12%+ and sub-1%, for various reasons like demography, healthcare readiness, inherent immunity, etc. ***Public perception has been created that fatality in India will be 3%-4%***. As already explained, this uses a flawed metric. The closest assumption to Fatality Rate in India is OFR as we have defined it, 9% at present.

OFR will India will decline further for two reasons, both related to low testing rates so far. One, because of low testing many late (advanced) cases are being diagnosed, who expire earlier than the assumed average 15 days from diagnosis. Thus the number of deaths in the OFR calculation is contaminated by numbers less than 15 days old, who are not part of the denominator. Second, as testing rates increase, they will identify more asymptomatic or mild cases, who will go through their 15-day disease process and survive – again lowering the OFR.

### Likely CFR in India and Forecasts

If OFR in India were to decline, what eventual figure (Case Fatality Rate) might it reach? Predicting this figure is out of the remit of mathematical modelling, and belongs at best to medical science. Irrespective of questionable data, China could be an analogy, since it is closest to India by demographic profile. China’s Fatality Rate presently announced is 5.5%**^(3)^**, and this is likely to stay that way since fresh cases are few.

We make the conscious assumption that India’s OFR, currently 9%, *will reduce by 1 percentage-point every week*, starting from Monday 11^th^ May, as testing approaches steady-state. **India’s CFR will eventually stabilise at 5.5% from 1^st^ June**.

Subject to the usual disclaimer, the model predicts that India will have **nearly 5000 deaths by May 31, may reach a total of 50000 deaths by mid-August and 100000 by mid-Sept**.

## D) EXPLORING ACTIVE CASES

Routinely announced measures of Recovery Rate are flawed for the same reason as Fatality Rates – they divide current Total Recoveries by current Total Infected cases. Since the mean time from diagnosis to recovery has been estimated as 15 days, the right metric is to divide Total Recoveries by Total Infected Cases 15 days ago. By including numbers diagnosed in the last 15 days, who have not yet lived out their disease (and add neither to Number of Recoveries or Deaths), we inflate the denominator and derive a Recovery Rate lower than what it actually is.

We define a term **Ongoing Recovery Rate (ORR)**, on day D being the Total Recoveries divided by the Total Infected Cases 15 days ago (D-15). In India, **ORR has remained steady, varying between 73.2% and 76.2% over Day 55 to Day 65, averaging 74.95%**. Eventually, at the end-point of the epidemic, when all cases have either recovered or expired, ORR will equal (1-CFR). Since we have assumed that CFR in India may be 5.5%, the final Recovery Rate (Case Recovery Rate) will be 94.5%.

The physical significance of these Ratios is expressed by the following statements:

- On any day D, the Total Number of Deaths will be OFR x Total Infected Cases on D-15.
- On any day D, the Total Recovered Cases will be ORR x Total Infected Cases on D-15
- Therefore, on any day D, *Total Active Cases* will be Total Infected Cases on day D, less Total Deaths on day D, less Total Recovered on day D.

Since the current ORR of 75% will eventually rise to 94.5% (1-CFR), for ease of computation we make the following assumptions -- ORR=75% till 10^th^ May, 80% from 11^th^ – 31^st^ May, 85% in June, and 90% thereafter.

Using the relations above, we computef Total Active Cases. **As shown in Fig 5, the forecasts are 88000 Active Cases by May-end, 200,000 by early July, and 400,000 by 18^th^ Aug**.

**Fig 5:**
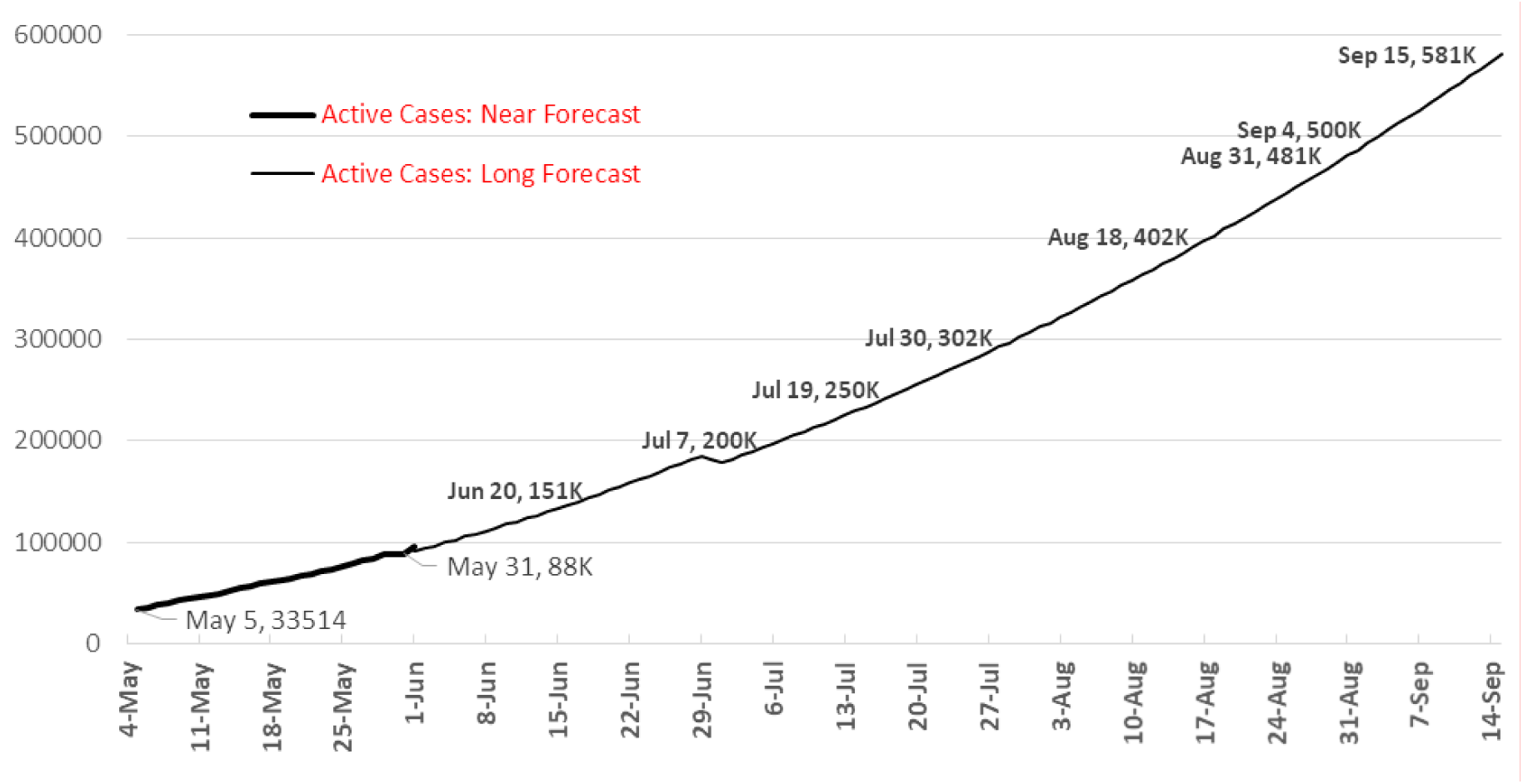
Total Active Cases in India, Short & Long Term Forecasts.

### Healthcare System Implications

*Indian Covid19 protocol currently requires all Covid-diagnosed patients to be in Hospital isolation*. Union Minister of Heath & Family Welfare Dr Harsh Vardhan stated**^(4)^** that there were 301634 dedicated Covid Hospital Beds in India. A newspaper article**^(5)^** reports that India has 400,000 isolation beds in hospitals. 50% of all available beds would be in use by early July, if current hospital protocol continues. Since hospital beds are location-specific, many regions would be near their maximum limits at this stage. We expect some policy changes by end-June. Either, self-isolation of mild cases without hospitalisation - this change would make availability of hospital facilities a non-issue till end-August. Also additional hospital facilities would be created. Creating the infrastructure for such facilities is not the problem -- the challenges here are the availability of doctors, para-medical staff, PPE, medicines and consumables. Implementation plans would have to begin by mid-April, if this option were to be chosen.

Another aspect of healthcare shortage – that of ICU beds and ventilators - is not explored as all available indications are that ventilator needs would be addressed by a combination of planned production/imports and innovative solutions from several agencies. It is assumed that hospital beds can be repurposed into ICUs if required.

**The conclusion from this exploration of active cases is that though hospital facilities are stretched by early July, hospital capacity is not an unsurmountable problem, if policy decisions are taken early and associated software issues planned for**.

The forecasts generated by the model in the foregoing sections are summarised in Table 2. 15^th^ Sept (4 months away) has been chosen as the end-date of forecasts on the assumption that some medical solution would become visible by then, even if not implemented.

**Table 2:** Forecast of key parameters till Sept 15 assuming model remains unaltered.

| By Date ... | Total Cases | Total Recoveries | Total Deaths | Total Active Cases | Doubling time at this stage | Remarks |
| --- | --- | --- | --- | --- | --- | --- |
| May 15<sup>th</sup> | 85000 | 28000 | 2500 | 54500 | 12 days | Near 1L Total Cases |
| May 31<sup>st</sup> | 169000 | 76000 | 5000 | 88000 | 16 days | Near 1L Active Cases |
| June 15<sup>th</sup> | 286000 | 144000 | 9000 | 133000 | 19 days | Near 10K Tot Deaths |
| June 30<sup>th</sup> | 448000 | 250000 | 16000 | 182000 | 20 days | Near 2L Active Case; Near 5L Total Cases |
| July 15<sup>th</sup> | 662000 | 404000 | 25000 | 233000 | 25 days | 25K deaths and near 250K Active Cases |
| July 31<sup>st</sup> | 954000 | 611000 | 37000 | 306000 | 30 days | Near 1 mn cases & 400K Active Cases |
| Aug 15<sup>th</sup> | 1.28 mn | 1.28 mn | 53000 | 386000 | 32 days | 50K deaths crossed 13<sup>th</sup> Aug |
| Aug 31<sup>st</sup> | 1.74 mn | 1.74 mn | 73000 | 481000 | 35 days |  |
| Sep 15<sup>th</sup> | 2.25 mn | 2.25 mn | 96000 | 581000 | 38 days | Near 1L Deaths |

## E) HOW FLAT IS FLAT?

The curve appears “flat enough” for healthcare infrastructure proposed, though shortage of people and consumables would remain a challenge.

*But the curve is NOT FLAT ENOUGH from the point of human cost – 100000 lives lost by the next 4 months, possibly the highest figure worldwide. Such heavy human casualties must be avoided*. **So how flat should, and can, the curve be?**

We set a limit of 50,000 deaths by Sept 15, and derive backwards the model of pandemic spread that reaches this figure, which by the by the current model is reached on 13^th^ Aug, That is, the proposed change delays the outcome by about 32 days. This slowing-down limits the spread after Sep 15^th^ as well.

To reduce the number of deaths reached by a given date, we can either reduce the Ongoing Fatality Rate (OFR), or the numbers infected. We have already factored-in OFR reduction to 5.5% from June 1, and there is no basis to assume further reductions Our option is to reduce the growth of numbers infected.

We make realistic assumptions about when one can implement policy interventions to bring growth *below present rate of growth*. Considering further relaxations after Lockdown 3.0 and massive inter-state migration and repatriation plans, it is unlikely that tougher regulations can come into effect before June 1. Thus our task is to find a pattern and growth trajectory of Total Infected Cases between Jun 1 and Sep 15 (106 days) that leads to a Total Deaths figure of 50,000 in contrast to the figure of nearly 100000 reached by the current trajectory.

The required model has been determined,. Fig 6 shows the graph of the current model, and the graph of the desired model, naturally much flatter. The current model has an overall doubling rate of 23 days in the period Jun 1 – Sep 15. In contrast, the desired model that limits fatalities should have a doubling rate of 38 days. **This will call for massive changes in social behaviour and regulations**. This study cannot speculate what such measures can be - mandated protective equipment, more severe containment strategies, modified forms of lockdown, or any other.

**Fig 6:**
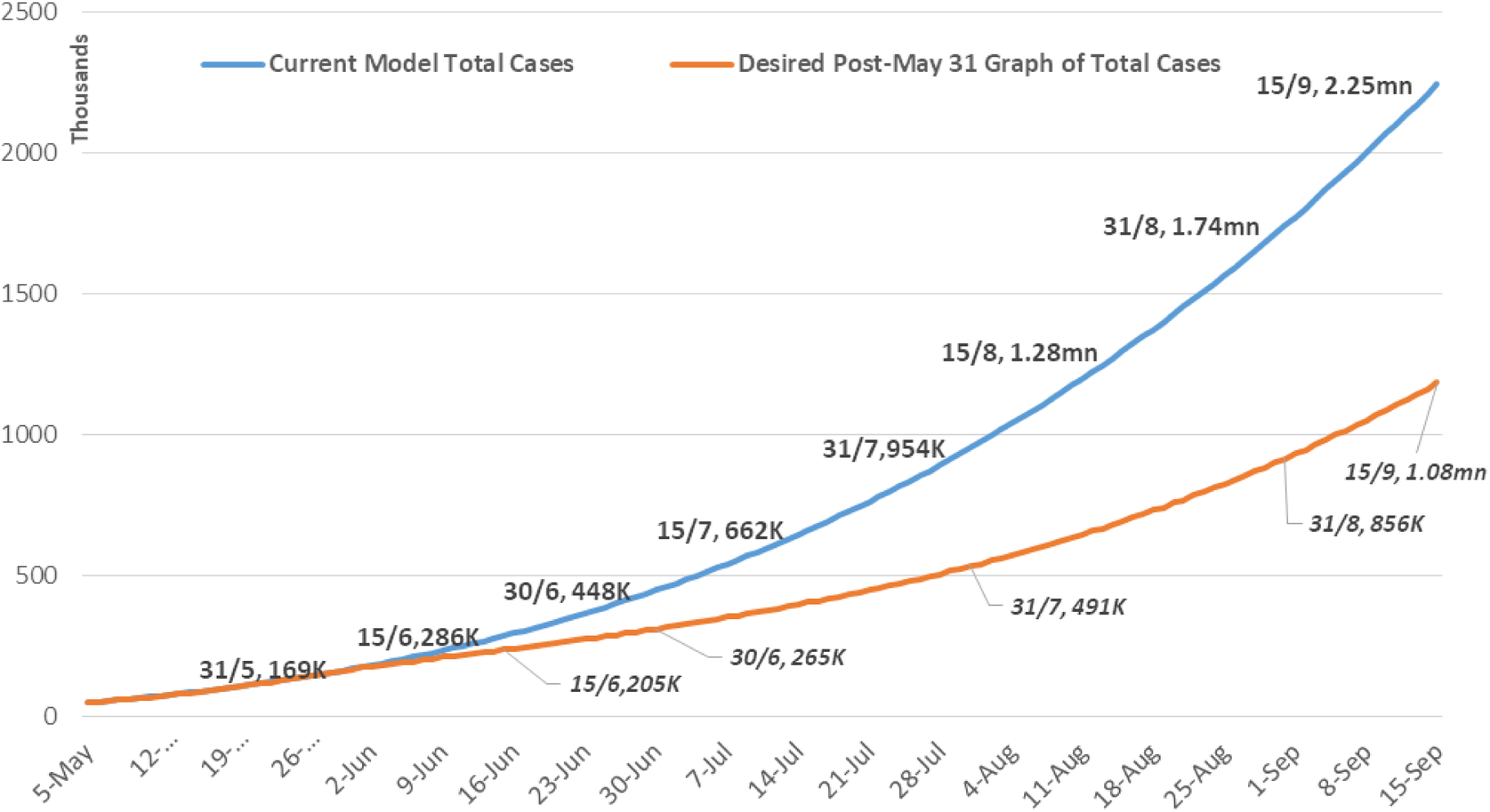
Current Predictor Curve vs Curve to Limit Deaths to 50K by 15 Sep.

Fig 7 reconfirms that Total Deaths would stay within our self-imposed limit by Sep 15 if the growth of Total Cases were as per the model in Fig 6. Recoveries and Active Cases can be computed as earlier, for the new desired model, and it is found that the Maximum Total Active Cases (on Sep 15) is 313,000, far lower than 581,000 going by the current curve.

Table 3 is the final summary that shows Total Cases, Total Deaths, Total Active Cases and Doubling Time under two situations. The first, the model that the growth of Total Cases has steadfastly followed for the last 30 days with exceptionally high R^2^. The second, the desired model that Total Cases **must follow** – in order that Total Deaths are limited to 50,000 by Sept 15^th^, with the added restriction that no change in pattern should be expected before June 1.

**Fig 7:**
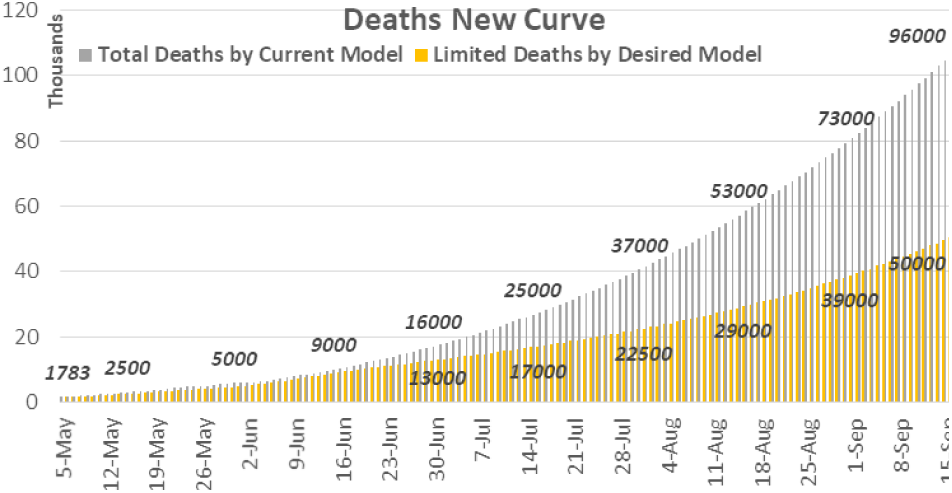
Total Deaths by Current Model vs Limited Deaths New Curve.

**Table 3:** Comparison of key parameters between Current and Desired Models.

| By Date ... | Situation 1: Current Predictor Curve |  |  |  | Situation 2: Desired Predictor Curve |  |  |  |
| --- | --- | --- | --- | --- | --- | --- | --- | --- |
|  | Total Cases | Total Deaths | Total Active Cases | Doubling time at this stage | Total Cases | Total Deaths | Total Active Cases | Doubling time at this stage |
| May 15<sup>th</sup> | 85000 | 2500 | 54500 | 12 days | 85000 | 2500 | 54500 | 12 days |
| May 31<sup>st</sup> | 169000 | 5000 | 88000 | 16 days | 169000 | 5000 | 88000 | 16 days |
| June 15<sup>th</sup> | 286000 | 9000 | 133000 | 19 days | 239000 | 9000 | 86000 | 24 days |
| June 30<sup>th</sup> | 448000 | 16000 | 182000 | 20 days | 311000 | 13000 | 89000 | 32 days |
| July 15<sup>th</sup> | 662000 | 25000 | 233000 | 25 days | 407000 | 17000 | 110000 | 37 days |
| July 31<sup>st</sup> | 954000 | 37000 | 306000 | 30 days | 536000 | 22500 | 144000 | 39 days |
| Aug 15<sup>th</sup> | 1.28 mn | 53000 | 386000 | 32 days | 693000 | 29000 | 181000 | 40 days |
| Aug 31<sup>st</sup> | 1.74 mn | 73000 | 481000 | 35 days | 935000 | 39000 | 240000 | 40 days |
| Sep 15<sup>th</sup> | 2.25 mn | 96000 | 581000 | 38 days | 1.19 mn | 50000 | 313000 | 40 days |

## F) CONCLUSION

At the end, we reiterate the objectives and results of this study. The study has presented the current model of Covid19 cases in India over time. Based on the stability of the model, while elaborating the events that would cause the model to change, the study has extrapolated Total Cases, Total Deaths and Active cases till Sept 15^th^. In doing so, the study has argued that the current publicly-reported Fatality and Recovery Rates are flawed, and derived more realistic estimates of these parameters. The forecasted numbers provide a basis for comparison with the results of planned policy interventions – if the actual observed data is less than the forecasts, we are doing well. This is the more important reason to use the forecasts, rather than as astrological predictions of the shape of things to come!

While exploring the number of Active Cases forecasted by the model, the study has found that Indian healthcare infrastructure has the capacity to deal with number of cases projected, more or less. In that sense, the Indian curve is flat enough. However, the current curve of Covid19 cases is NOT flat enough with respect to expected fatalities, which by the current growth curve would reach 100,000 by Sept 15^th^. The study set an arbitrary limit of 50000 Total Deaths by Sep 15, and retro-fitted a mathematical model that was acceptably flat. The key parameters of this desired curve – Total Cases, Total Deaths and Active Cases – have been tabulated fortnightly, as also done for the current model.

While we look forward to Public health initiatives to tackle the pandemic, the outputs of this study provide the reference points against which the success or failure of public health initiatives can be judged.

## Data Availability

All data reported in the manuscript are available in the public domain. All significant references have been cited.

https://www.ndtv.com/coronavirus/india-covid-19-tracker

https://www.worldometers.info/coronavirus/coronavirus-death-rate/

